# Early treatment with nitazoxanide prevents worsening of mild and moderate COVID-19 and subsequent hospitalization

**DOI:** 10.1101/2021.04.19.21255441

**Authors:** Jean-François Rossignol, C. Bardin Matthew, Joshua B. Oaks, B. Gregory Bostick, Kishor N. Vora, Jessica Fulgencio, Dena Mogelnicki, Christian Bréchot, Vanguard Study Group

**Author notes:** Members of the study group are listed in the Appendix (Vanguard Study Group).

## Abstract

**Background:** There is an urgent need for treatments to prevent the progression to severe COVID-19 and hospitalization.

**Methods:** A randomized double-blind placebo-controlled clinical trial in 36 centers in the U.S. and Puerto Rico investigated the safety and effectiveness of oral nitazoxanide 600 mg twice daily for 5 days in outpatients with symptoms of mild or moderate COVID-19 enrolled within 72 hours of symptom onset. Key objectives were reduction of duration of symptoms (primary) and progression to severe illness (key secondary).

**Results:** 1,092 subjects were enrolled, and 379 with laboratory-confirmed SARS-CoV-2 infection were analyzed. Overall, times to sustained clinical recovery were similar for the two arms. Nitazoxanide treatment was associated with an 85% reduction in the progression to severe COVID-19 (1/184, [0.5%] vs. 7/195, [3.6%], p=0.07) and 82% reduction in the rate of hospitalization, emergency room visit or death (1/184 [0.5%] vs. 6/195 [3.1%], p=0.12). In subjects with mild illness at baseline, treatment was also associated with a 3.1-day reduction in median time to sustained clinical recovery and a 5.2-day reduction in time to return to usual health. Nitazoxanide was safe and well tolerated.

**Conclusions:** Treatment of mild or moderate COVID-19 with a five-day course of oral nitazoxanide was safe and well tolerated and was associated with an 85% reduction in the progression to severe illness and a 3- to 5-day reduction of the duration of mild illness.

## INTRODUCTION

The progress in developing effective vaccines against SARS-CoV-2 infection has been remarkable. Coupling effective therapies with vaccination programs will be critical to controlling the pandemic.^1^ However, the outcome of significant efforts to develop novel therapies, particularly in outpatients, has so far been limited. Promising results have been achieved with monoclonal antibodies, yet these treatments are costly, prone to viral resistance, and can only be administered in a clinic setting^2–4^. Hence, there is still a need for medications which can be distributed broadly and in settings with scarce healthcare resources to prevent the worsening of mild or moderate COVID-19 symptoms and subsequent hospitalization.

Nitazoxanide is approved for use in the United States for the treatment of diarrhea caused by *Cryptosporidium parvum* and *Giardia intestinalis* infections and has been used throughout Latin America and Asia for the treatment of intestinal parasitic infections. In the 25 years since nitazoxanide was first introduced, approximately 500 million people have been treated worldwide, and the drug has demonstrated a favorable safety record in both adults and children. Nitazoxanide has been previously shown to be active *in vitro* against a broad range of viruses including MERS and certain animal coronaviruses^5–7^, as well as to suppress secretion of cytokines associated with the inflammatory response to respiratory infection^8,9^. Recently, nitazoxanide was identified as a candidate drug for SARS-CoV-2 infection based on high throughput screening and *in vitro* virus culture^10–14^.

The antiviral activity of nitazoxanide is attributed to a host-directed mechanism, ultimately targeting the formation of key viral proteins at a post-translational level^15,16^. Due to this mechanism, studies with other viruses suggest nitazoxanide has a low potential to develop resistance^17,18^. Furthermore, multiple studies indicate nitazoxanide has a synergistic effect when combined with other drugs active against SARS-CoV-2^12,14,19^.

A recent study has demonstrated *in vitro* that nitazoxanide blocks the spike maturation of the B.1.1.7 and P1 mutants with the same efficacy as for the reference Wuhan strain and D614G mutations^11^.

In a multicenter, randomized, double-blind, placebo-controlled trial in patients hospitalized with moderate-to-severe COVID-19, treatment with nitazoxanide 600 mg twice daily for seven days was associated with reductions in rates of mortality and mechanical ventilation, duration of supplemental oxygen and time to hospital discharge compared to placebo^20^.

The present multicenter, randomized, double-blind, placebo-controlled trial provides evidence nitazoxanide prevents the progression to severe illness and hospitalization and reduces the duration of mild illness when administered to patients within 72 hours of symptomatic SARS-CoV-2 infection.

## METHODS

### Study design and subjects

This was a randomized double-blind placebo-controlled trial conducted in 36 outpatient medical clinics in the U.S. and Puerto Rico in accordance with current good clinical practices and applicable regulations (ClinicalTrials.gov Identifier: NCT04486313).

Subjects at least 12 years of age presenting within 72 hours of onset of symptoms of mild or moderate COVID-19 were eligible to participate in the trial. Minimum symptom requirements were: at least two respiratory symptom domains (head, throat, nose, chest, cough) with a score of ≥2 as determined by scoring the InFLUenza Patient-Reported Outcomes (FLU-PRO^©^)^21^ questionnaire administered at screening (only one domain score required to be ≥2 if pulse rate ≥90 beats per minute or respiratory rate ≥16 breaths per minute), with no improvement in overall symptom severity from the prior day. Key exclusion criteria were: (i) signs or symptoms suggestive of severe COVID-19 including shortness of breath at rest, resting pulse rate ≥125 beats per minute, resting respiratory rate ≥30 breaths per minute, or SpO2 ≤ 93% on room air at sea level; (ii) previous COVID-19 infection, (iii) immunodeficiency; and (iv) pregnant females and sexually active females of childbearing potential not using birth control.

Nitazoxanide was administered as two 300 mg extended release tablets (600 mg per dose) orally with food twice daily for five days. The dose was selected based upon a dose-range-finding study in patients with influenza^22^.

### Randomization and masking

Eligible subjects were centrally randomized using an interactive web response system 1:1 to receive treatment with nitazoxanide or matching placebo tablets. In addition to study medication, all subjects received a vitamin B complex supplement (Super B-Complex(tm), Igennus Healthcare Nutrition, Cambridge, UK) twice daily to mask any potential chromaturia attributed to nitazoxanide. The randomization list was masked to study participants, the sponsor, investigators, study monitors, and laboratory personnel until the database was locked.

Randomization was stratified according to the severity of COVID-19 illness at baseline (mild or moderate), time from onset of symptoms (<36 hours or ≥36 hours), and whether subjects had risk factors for severe illness based on CDC criteria current at the initiation of the study (see Supplementary Material for CDC criteria). Moderate illness was defined by resting pulse ≥90 beats per minute and/or resting respiratory rate ≥20 breaths per minute.

### Study procedures

After randomization, eligible subjects underwent a physical examination, collection of nasopharyngeal swabs, and blood and urine samples for laboratory safety testing and assessment of SARS-CoV-2 antibody titers. Study drug was dispensed, and subjects were followed for 28 days. Subjects were instructed to complete electronic diaries recording oral temperature twice daily and symptom severity once daily in the evening for 21 days and were contacted daily by telephone by site staff on study days 2-7 and 28 to verify compliance and screen for progression to severe illness or other complications. Repeat nasopharyngeal swabs were collected on study days 4 and 10. Follow up blood and urine samples for laboratory safety testing and assessment of SARS-CoV-2 antibody titers were collected on study day 22. Subjects were allowed to use acetaminophen and/or dextromethorphan for symptom relief along with standard-of-care rescue medications. Adverse events were collected continuously throughout the study and monitored until the events resolved.

In the absence of any patient-reported outcomes instrument validated specifically for collecting data to measure symptoms of COVID-19, symptom data was collected using the FLU-PRO Plus^©^ symptoms questionnaire. The InFLUenza Patient-Reported Outcome Questionnaire (FLU-PRO^©^) was developed in accordance with psychometric best practices and FDA guidance for the measurement of symptoms of influenza^21^. Subsequent literature searches and clinical data analyses support the content and construct validity of the instrument for illness caused by non-influenza respiratory viruses including adenovirus, endemic coronaviruses, enteroviruses including rhinoviruses, parainfluenza and respiratory syncytial virus. A literature search conducted prior to initiation of the study supported content validity of the FLU-PRO Plus^©^ with inclusion of loss of taste and loss of smell for measurement of illness caused by SARS-CoV-2 infection. The questionnaire was completed using an electronic diary app downloaded to each subject’s smart phone or a provisioned electronic device so that diary entries were time stamped to ensure timely recording, thereby mitigating risks of recall bias.

Nasopharyngeal swab samples collected at baseline, day 4 and day 10 were tested using the Aptima^®^ SARS-CoV-2 assay (Hologic, Inc, San Diego, CA) and ePlex^®^ Respiratory Pathogen Panel (“ePlex RPP”, GenMark, Carlsbad, California). Baseline, day 4 and day 10 nasopharyngeal swab samples positive for SARS-CoV-2 by the Aptima^®^ SARS-CoV-2 assay were subjected to RT-PCR for analysis of quantitative changes in viral load. Blood samples collected at baseline and day 22 were tested for quantitative anti-SARS-CoV-2 antibodies.

### Primary and Secondary Outcomes

The primary endpoint was time from the first dose to sustained response (TSR), a measure of meaningful within-subject symptom improvement developed and validated in subjects with influenza infection. The performance characteristics of the FLU-PRO instrument and appropriateness of background levels in subjects with SARS-CoV-2 infection were confirmed by blinded analysis of diary data for this study after database lock and prior to unblinding.

The key secondary endpoint was the rate of progression to severe COVID-19 illness (shortness of breath at rest and SpO_2_ ≤ 93% on room air or PaO_2_/FiO_2_ <300). This definition was selected over a definition including hospitalization due to variability in physician decisions regarding hospital admission.

### Statistical analysis

Efficacy analyses were based on a modified intention to treat (mITT) population of subjects testing positive for SARS-CoV-2 at baseline. All subjects receiving at least one dose of study medication were included in the safety analyses.

In the primary analysis, TSR for the nitazoxanide treatment group was compared to that of the placebo treatment group using a stratified Gehan-Wilcoxon test (α=0.05) where stratification followed that used for randomization. Subjects without a sustained response recorded were treated as censored as of the last diary completed, except for subjects who were hospitalized or died during the study, who were censored at Day 21.

In the absence of prior experience in subjects with COVID-19, the sample size was determined based upon data from two prior clinical trials of nitazoxanide in subjects with viral respiratory illnesses caused by influenza or rhinoviruses. A sample size of 312 subjects (156 per group) was calculated to provide 90% power to detect a statistically significant difference in the survival distributions between the nitazoxanide and placebo groups (Gehan rank test, two-sided α =0.05).

In the key secondary analysis, proportions of subjects progressing to severe COVID-19 illness were compared between the treatment groups using a Cochran-Mantel-Haenszel (CMH) test stratified by the randomization strata.

## RESULTS

From August 18, 2020 through January 8, 2021, 1,092 subjects were enrolled at 36 sites in the U.S. and Puerto Rico, including 379 with laboratory-confirmed SARS-CoV-2 infection (**Figure 1**). Demographic and disease-related characteristics of the SARS-CoV-2-infected population are summarized in **Table 1**.

**Table 1:** **Summary of Baseline Demographic and Disease-Related Characteristics, ITTI Population**

|  | **Nitazoxanide**<br>**(N=184)** | **Placebo**<br>**(N=195)** | **All mITT**<br>**Subjects**<br>**(N=379)** |
| --- | --- | --- | --- |
| Male, N (%) | 83 (45.1%) | 82 (42.1%) | 165 (43.5%) |
| Median (Range) Age (years) | 38 (12-83) | 42 (13-81) | 40 (12-83) |
| Race or Ethnic Group, N (%) |  |  |  |
| White | 117 (63.6%) | 116 (59.5%) | 233 (61.5%) |
| Hispanic or Latino | 59 (32.1%) | 71 (36.4%) | 130 (34.3%) |
| Black or African American | 4 (2.2%) | 4 (2.1%) | 8 (2.1%) |
| Asian | 2 (1.1%) | 4 (2.1%) | 6 (1.6%) |
| Other | 2 (1.1%) | 0 (0.0%) | 2 (0.5%) |
| Median (IQR) BMI | 28.7 (25.3, 33.5) | 29.1 (25.7, 33.6) | 28.9 (25.5, 33.5) |
| Median Hours Since Onset of Symptoms | 43.9 | 46.5 | 45.9 |
| Moderate Disease Severity, N (%) | 68 (37.0%) | 65 (33.3%) | 133 (35.2%) |
| At Risk of Severe Illness per CDC Guidelines, N (%) | 112 (60.9%) | 126 (64.6%) | 238 (62.8%) |
| Viral Load, log<sub>10</sub> RNA copies/mL |  |  |  |
| Median (IQR) | 6.38 (4.71, 7.40<sup>1</sup>) | 6.34 (4.40, 7.40<sup>1</sup>) | 6.36 (4.62, 7.40<sup>1</sup>) |
| SARS-CoV-2 Antibody-Positive, N (%) | 18 (4.7%) | 20 (5.3%) | 38 (10.0%) |

<sup>1</sup> 7.40 log<sub>10</sub> copies/mL is the upper limit of quantitation of the assay.

**Figure 1.**
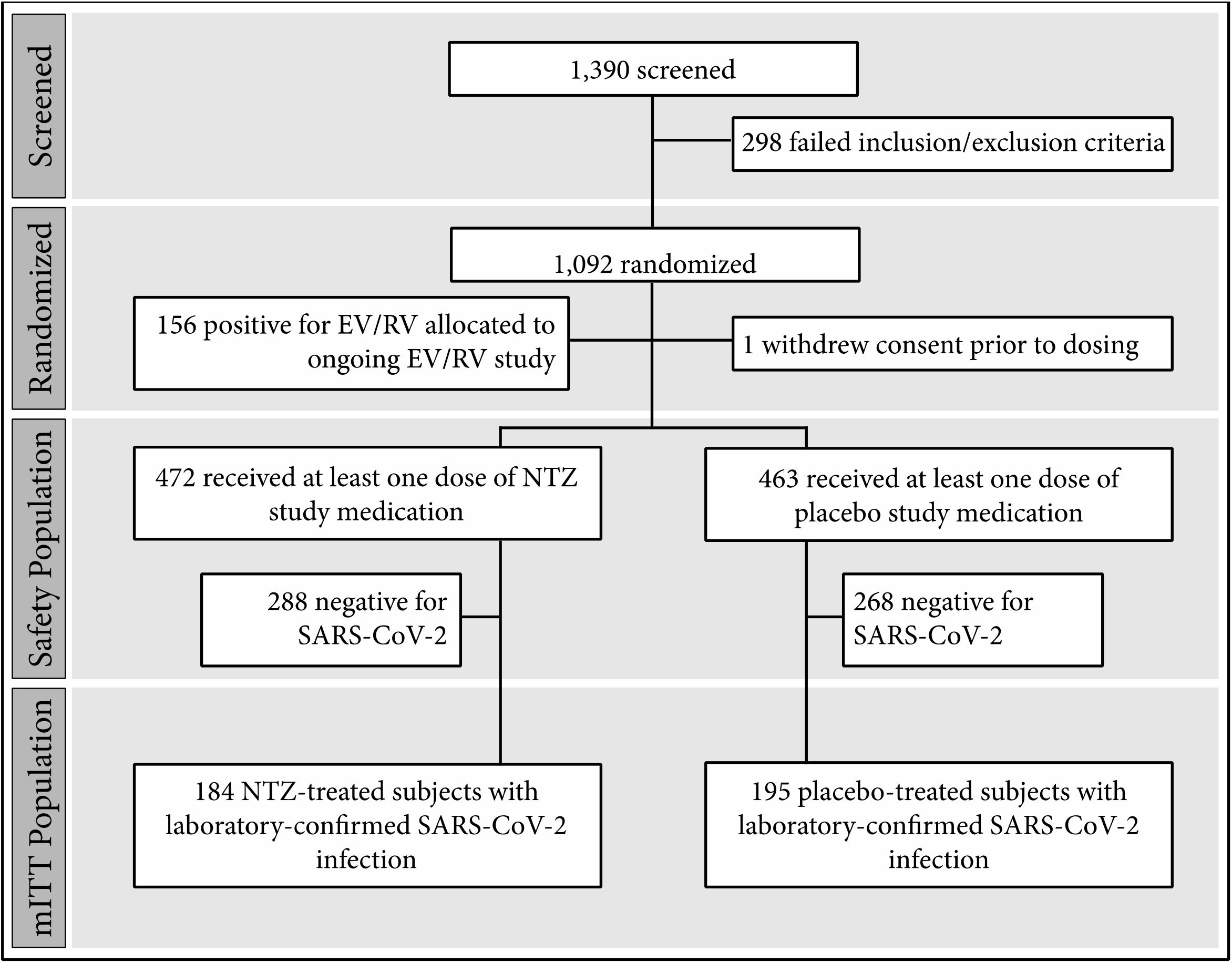
Subject Disposition.

### Primary Outcome

Ninety-two percent (92%) of all possible daily FLU-PRO questionnaires were completed, and only 22 (5.8%) subjects were censored in the primary analysis due to missing diary data prior to the day 22 visit. Median (IQR) TSR were 13.28 (6.26 - >21) and 12.35 (7.18 - >21) days for the nitazoxanide and placebo groups, respectively (p=0.88).

### Key Secondary Outcome

Eight subjects met the criteria for progression to severe COVID-19, **Table 2**. Treatment with nitazoxanide was associated with an 85% reduction of progression to severe illness compared to placebo (1/184 [0.5%] for the nitazoxanide group compared to 7/195 [3.6%] for the placebo group, p= 0.07), **Table 3**.

**Table 2:** Subjects Progressing to Severe COVID-19 Illness.

| **Treatment Group** | **Age Group** | **Sex** | **BMI** | **Viral Load<sup>1</sup>**<br>**Day 1/Day 4** | **Comorbidities** | **Study Day** | **Medical Attention Required** | **Secondary Diagnosis** | **SpO<sub>2</sub>** |
| --- | --- | --- | --- | --- | --- | --- | --- | --- | --- |
| Placebo | 70-80 | M | 31.7 | 6.18 / $\geq 7.40$ | Type 2 diabetes mellitus, Parkinson's disease, neuropathy, colon cancer/colon re-sectioning, benign prostatic hyperplasia, obesity | 7 | Hospitalized | COVID-19 pneumonia | 88% |
| NTZ | 70-80 | M | 25.5 | 7.39 / 4.79 | Hypertension, coronary artery disease, atrial fibrillation, bilateral iliac stents, type 2 diabetes mellitus, peripheral vascular disease, hyperlipidemia, decreased renal function, cholecystectomy, prostate cancer with prostatectomy, hypogonadism, gout | 5 | Hospitalized | COVID-19 pneumonia | 91% |
| Placebo | 50-60 | F | 22.8 | 5.96 / $\geq 7.40$ | Hypertension, anxiety, migraine headaches, cigarette use | 3 | Hospitalized | Syncope | 93% |
| Placebo | 60-70 | F | 36.4 | 6.54 / 7.06 | Hyperlipidemia, osteoarthritis, insomnia, anxiety, obesity | 10 | Emergency room | COVID-19 pneumonia | 93% |
| Placebo | 60-70 | M | 33.0 | ND / ND | Hypertension, hypercholesterolemia, pre-diabetes, gastroesophageal reflux disease, osteoarthritis, anxiety, prostate cancer and surgery, bilateral hip arthroplasty, cataracts, obesity | 7 | Hospitalized | Dyspnea | 88% |
| Placebo | 50-60 | M | 26.8 | $\geq 7.40$ / NC | Type 2 diabetes mellitus, borderline reduced kidney function, coronary artery disease, coronary artery bypass graft, neuropathy, hypothyroidism, hernias | 9 | Hospitalized | COVID-19 pneumonia | 84% |
| Placebo | 30-40 | M | 35.4 | $\geq 7.40$ / IND | Essential hypertension, elevated liver enzymes, asthma, obesity | 5 | Clinic visit | Asthma (exacerbation) | 93% |
| Placebo | 50-60 | M | 34.9 | 6.47 / ND | Hypertension, elevated liver enzymes, previous stroke, subcarinal adenopathy, migraines, nerve issues due to injury, obesity | 7 | Hospitalized | COVID-19 pneumonia | 90% |

<sup>1</sup>log<sub>10</sub> RNA copies/mL, upper limit of quantitation of the assay was 7.40 log<sub>10</sub> RNA copies/mL
<sup>2</sup>ND = Not Done due to insufficient sample; NC = Sample Not Collected (visit missed), IND = Indeterminate result by RT-PCR

**Table 3:** **Analyses of the Subjects Progressing to Severe COVID-19 with Subgroups Based Upon Different At-Risk Definitions**

|  | Placebo | NTZ | Relative Reduction |
| --- | --- | --- | --- |
| All SARS-CoV-2-Positive Subjects | 7/195 (3.6%) | 1/184 (0.5%) | 85% |
| Subgroups: |  |  |  |
| “May Be” or “At Increased Risk” for Severe COVID-19 Illness per CDC (protocol-defined stratification factor) | 7/126 (5.6%) | 1/112 (0.9%) | 84% |
| At Increased Risk for Severe COVID-19 Illness per CDC | 7/121 (5.8%) | 1/104 (1.0%) | 83% |
| At High Risk of Progressing to Severe COVID-19 and/or Hospitalization (per EUA documents for monoclonal antibodies)<sup>1</sup> | 6/69 (8.7%) | 1/60 (1.7%) | 81% |

<sup>1</sup> ≥65 years of age, BMI ≥35 kg/m<sup>2</sup>, chronic kidney disease, diabetes, immunosuppressive disease, current receipt of immunosuppressive treatment, or ≥55 years of age with at least one of cardiovascular disease, hypertension, or chronic obstructive pulmonary disease or another chronic respiratory disease

### Exploratory Outcomes

Treatment with nitazoxanide was associated with an 82% reduction in the rate of hospitalization, emergency room visit or death (p = 0.11), **Table 4**.

**Table 4:**
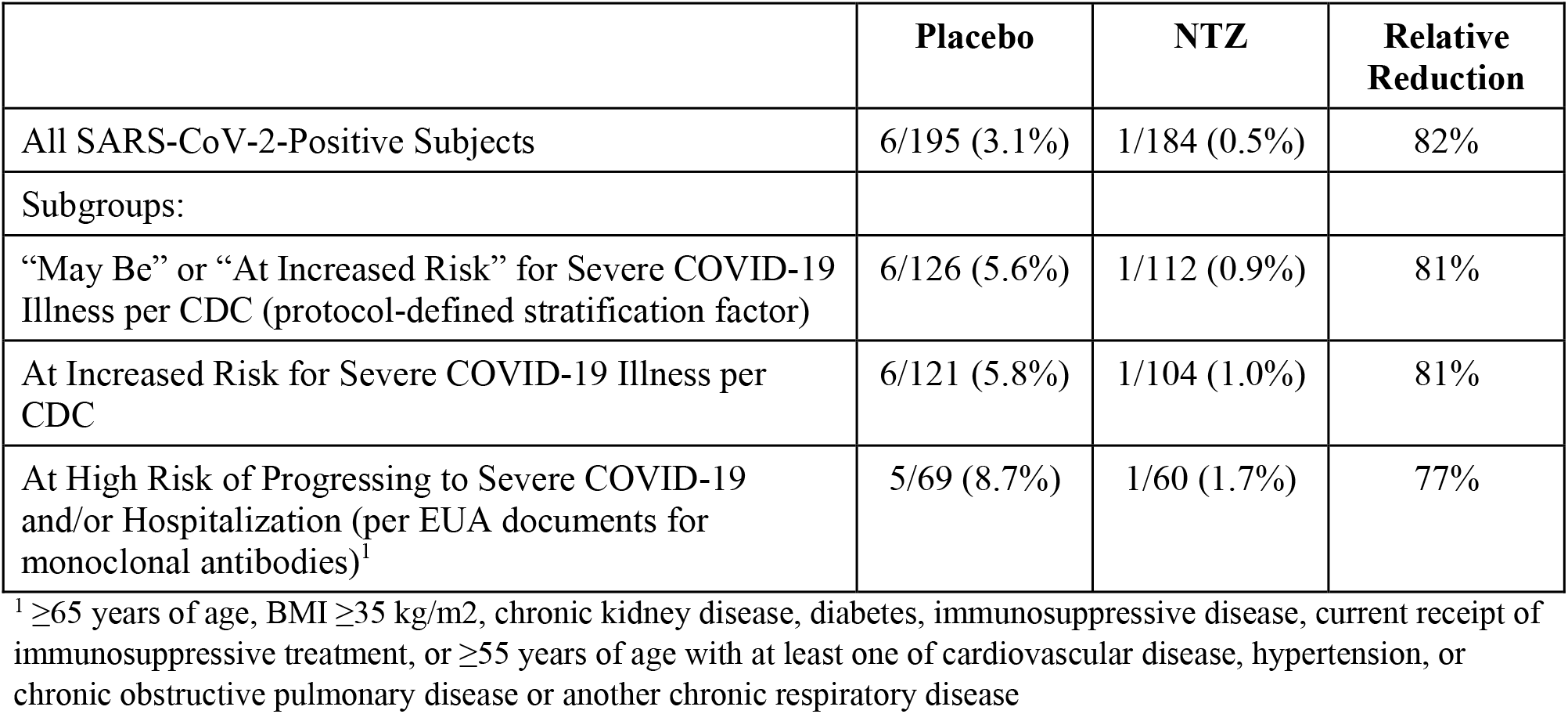
**Analyses of the Subjects Experiencing Hospitalization, Emergency Room Visit or Death with Subgroups Based Upon Different At-Risk Definitions**

Ninety four percent (94%) and 70% of subjects tested positive for SARS-CoV-2 RNA in nasopharyngeal swabs collected at study days 4 and 10, respectively. Qualitative and quantitative tests to detect SARS-CoV-2 were not significantly different between the treatment arms at these time points, **Table 5**.

**Table 5:** Virologic Data by Time Point.

| **Time Point** | **NTZ** |  |  | **Placebo** |  |  |
| --- | --- | --- | --- | --- | --- | --- |
|  | **N Samples Analyzed** | **Percent Positive** | **Mean (SD) Viral Load Change from Baseline (log<sub>10</sub> copies/mL)** | **N Samples Analyzed** | **Percent Positive** | **Mean (SD) Viral Load Change from Baseline (log<sub>10</sub> copies/mL)** |
| Day 4 | 177 | 94% | -0.70 (1.573) | 178 | 94% | -1.02 (1.668) |
| Day 10 | 177 | 73% | -2.49 (1.582) | 182 | 71% | -2.61 (1.604) |

In the 246 subjects (65%) with mild illness at baseline, treatment with nitazoxanide was associated with a 3.1-day reduction of median TSR (median [IQR] = 10.3 days [6.2->21] for nitazoxanide [n=116] compared to 13.4 days [7.4->21] for the placebo group [n=130], p= 0.0932) and a 5.2-day reduction of median time from first dose until the subjects reported returning to usual health (median [IQR] = 13.2 days [9.2->21] for nitazoxanide compared to 18.4 days [11.4->21] for the placebo group, p= 0.0075), **Figure 2**. In the 133 subjects (35%) with moderate illness at baseline, treatment with nitazoxanide was associated with a longer TSR and time to return to usual health. None of the 68 subjects with moderate illness in the nitazoxanide arm experienced progression to severe illness while two of 65 in the placebo arm did.

**Figure 2.**
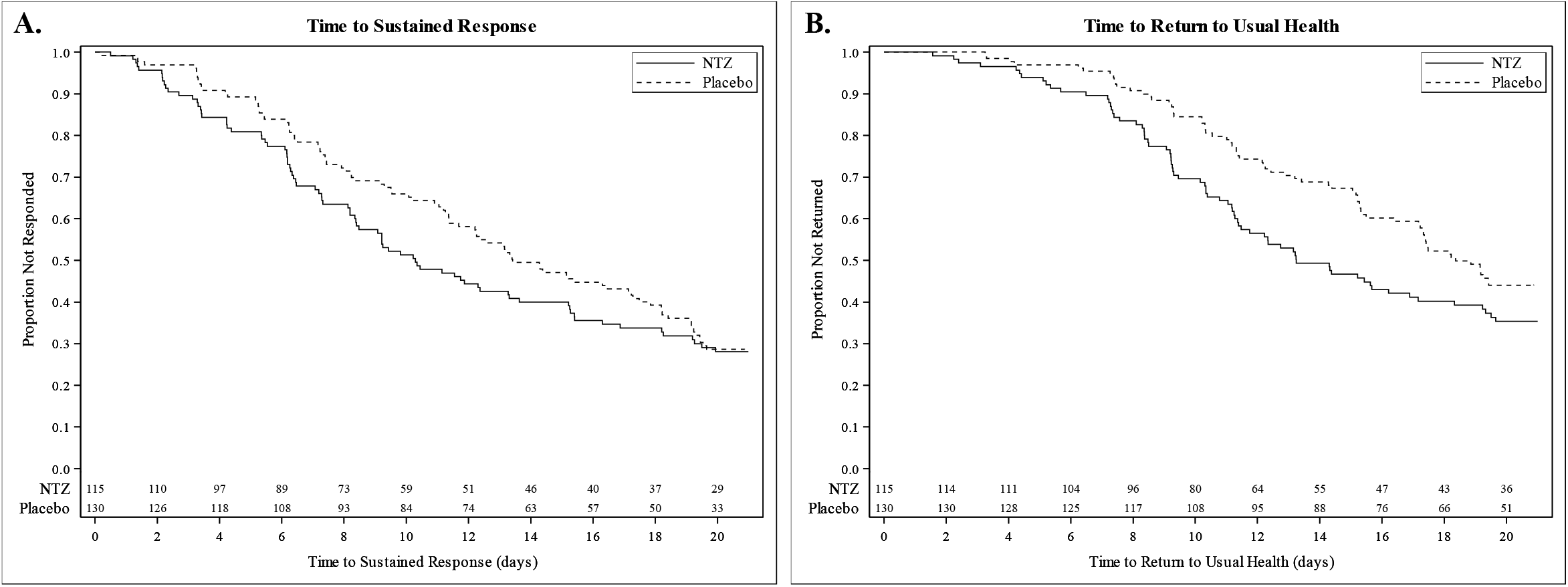
Clinical Recovery in Subjects with Mild Illness. (A) Time to sustained response in subjects with mild illness; (B) Time to return to usual health in subjects with mild illness

### Safety

Nitazoxanide was safe and well tolerated. Sixty-three subjects (13.3%) in the nitazoxanide group and 75 (16.2%) in the placebo group reported at least one adverse event, predominately classified as mild or moderate in severity and unrelated or possibly related to study drug. Only diarrhea was reported in ≥2% in any treatment group (n=16 [3.4%] in the nitazoxanide group and n=10 [2.2%] in the placebo group). Frequency, severity and assessment of relationship to study drug of adverse events were similar across treatment groups. Two subjects treated with nitazoxanide and seven receiving placebo reported serious adverse events, all unrelated to the study drug. Two subjects (both in the nitazoxanide treatment group) died during the study, one due to severe COVID-19 and the other (SARS-CoV-2 negative) secondary to aspiration 19 days after completing therapy. Neither event was considered related to study medication in the judgment of the Investigator, Sponsor, Medical Monitor and Independent Data Monitoring Committee. Two nitazoxanide-treated subjects and three receiving placebo discontinued study medication due to adverse events.

## DISCUSSION

The COVID-19 pandemic continues with surges caused by SARS-CoV-2 variants, and the rate of hospitalization poses a major threat to health care systems in many countries. Despite the development of vaccines which are now being distributed, widespread vaccination will need time to be implemented worldwide and will not fully prevent infection. Thus, there is a critical need for a safe, easy-to-administer antiviral therapeutic that can be distributed through pharmacies and administered early for treatment of mild or moderate COVID-19 – ideally a host-directed antiviral with a high barrier to resistance that could provide a line of defense against emerging variants.

We report a multicenter randomized double-blind placebo-controlled trial conducted at 36 outpatient centers in the United States and Puerto Rico between August 2020 and February 2021. The study employed a concurrent placebo control and enrolled a broad range of subjects at least 12 years of age, 63% of whom had risk factors placing them at higher risk of severe COVID-19. Subjects were enrolled based upon symptoms to ensure early treatment, avoiding limitations associated with the availability of and delays in diagnostic testing, and 379 subjects with confirmed SARS-CoV-2 infection were analyzed for effectiveness. The trial was appropriately blinded, and subjects were closely followed for 28 days. The trial was designed early during the course of the pandemic without the benefit of prior experience with COVID-19, nevertheless, the endpoints were objective, relevant and well-defined, and rigorous data collection procedures were employed.

Treatment with nitazoxanide 600 mg orally twice daily for five days was associated with an 85% reduction in the rate of progression to severe illness (7/195 vs. 1/184, p=0.07). All severe illnesses were clinically meaningful with six of the eight requiring hospitalization. Each of the eight severe illnesses occurred between study days 3 and 10 in subjects at high risk of severe illness according to CDC criteria. While the numbers of events observed are low, they are quantitatively and qualitatively similar or superior to those used to support emergency use of the first monoclonal antibodies for treating mild or moderate COVID-19 in the United States^2,3,23,24^.

These findings are supported by another multicenter, randomized, double-blind, placebo-controlled study recently reported by Blum et al. in subjects hospitalized with moderate to severe COVID-19 where treatment with nitazoxanide 600 mg twice daily for seven days was associated with reductions in rates of mortality and mechanical ventilation, duration of supplemental oxygen and time to hospital discharge compared to placebo^20^.

There is presently no consensus with respect to a symptoms-based endpoint for use in trials of therapeutics in subjects with mild or moderate COVID-19. For this trial, we used the FLU-PRO Plus questionnaire to collect symptom data over 21 days and an endpoint defined based upon evidence that it is meaningful to patients. Treatment with nitazoxanide was associated with a three- to five-day reduction of the duration of illness in subjects with mild illness at baseline, but not in those with moderate illness. Given present knowledge of the stages/phases of COVID-19 disease and their relevance to treatment decisions, it is reasonable that patients with mild illness may achieve full recovery more rapidly following an effective treatment than those with moderate illness. The subgroup with mild illness at baseline was a larger and more homogenous population and likely associated with less variability in time to full recovery than subjects with moderate illness. We note that none of the nitazoxanide-treated subjects with moderate illness at baseline progressed to severe illness compared to two subjects receiving placebo.

We did not observe differences between treatment groups in qualitative or quantitative SARS-CoV-2 RNA in nasopharyngeal swabs on study day 4 or 10. While others have reported modest reductions of quantitative or qualitative RNA in nasopharyngeal swabs at different points after the end of treatment with nitazoxanide,^20,25,26^ the methods used for collection, sample handling and measuring viral loads from nasopharyngeal swabs in large multi-center clinical trials have not been validated or shown to be predictive at the patient- or trial-level of viral load, inflammation or symptoms in the lungs or clinical outcomes. It is also unclear whether RT-PCR accurately measures infectious virus, since viral RNA may persist for some time, even in the absence of replication-competent virus. This may be a particularly important limitation in the context of a host-directed therapeutic like nitazoxanide that affects assembly of the virus.

In this study, nitazoxanide was safe and well tolerated, consistent with its well-established safety profile. Safety will be an important attribute for a therapeutic for mild or moderate COVID-19.

Preventing the progression to severe COVID-19 illness is a key factor for controlling the pandemic. Only monoclonal antibodies directed to the viral spike protein have shown promise when used at the early stage of infection, yet the emergence of SARS-CoV-2 resistance to these antibodies has already been observed, necessitating the development of antibody cocktails or rescission of emergency use authorization^2–4^. In this study, nitazoxanide reduced the progression to severe COVID-19 and hospitalization in a manner similar to that of the monoclonal antibodies.

Persons in rural and/or economically disadvantaged communities as well as racial and ethnic minorities in the U.S. and worldwide have been disproportionately impacted by the COVID-19 pandemic for numerous reasons. An oral shelf-stable drug for the treatment of COVID-19 that can be administered at home in these vulnerable populations may mitigate the burden of reduced access to healthcare resources such as an infusion center or pharmacy with cold storage that is necessary for administration of monoclonal antibodies.

The evidence provided by this study is meaningful, reliable and consistent with expectations of a therapy that may provide potential benefit to patients at high risk of progression to severe COVID-19 and/or hospitalization. Nonetheless, these results should be confirmed with larger trials. The availability of a safe, oral, scalable, host-directed antiviral for the early treatment of COVID-19 could play an important role in reducing the number of severe illnesses and hospitalizations during this ongoing major public health crisis.

## Data Availability

Reasonable requests from qualified researchers will be considered for data sharing. These requests should be submitted to.

## Acknowledgments

We thank the clinical centers which have contributed to this study.

## Appendix

### Vanguard Study Team (RM08-3008)

1. Maher Agha, MD (OnSite Clinical Solutions – Charlotte, Charlotte, NC)
2. Ayoade Akere, MD (Eagle Clinical Research, Chicago, IL)
3. Ali Bajwa, MD (Centex Studies – Westfield, Houston, TX)
4. Greg Bostick, MD (Cullman Clinical Trials, Cullman, AL)
5. Jose F. Cardona, MD (Indago Research & Health Center, Inc., Hialeah, FL)
6. Ivan Carreras, MD (Clintex Research Group, Inc, Coral Gables, FL)
7. Jorge Diaz, DO (Doral Medical Research, Inc., Hialeah, FL)
8. Dina Doolin, DO (Riverside Clinical Research, Edgewater, FL)
9. Timothy Elder, MD (SIMEDHealth, LLC, Gainesville, FL)
10. Almena L. Free, MD (Pinnacle Research Group, LLC, Anniston, AL)
11. Bernard Garcia, MD (Invesclinic U.S., Ft. Lauderdale, FL)
12. Hiram Garcia, MD (Rio Grande Valley Clinical Research Institute, Pharr, TX)
13. Darin M. Gregory, MD (Pioneer Clinical Research, Bellevue, NE)
14. Barry Heller, MD (Long Beach Clinical Trials, Long Beach, CA)
15. Rubaba Hussain, MD (Prime Global Research, Bronx, NY)
16. Talal Khader, MD (Vida Clinical Studies, Dearborn, MI)
17. Rogelio Machuca, MD (Machuca Family Medicine, Las Vegas, NV)
18. Eric J. Melvin, MD (Clinical Trials of America, LLC, Mt. Airy, NC)
19. Randall P. Miller, MD (Horizon Research Group of Opelousas, LLC, Eunice, LA)
20. Nidal Morrar, MD (G & L Research, Foley, AL)
21. Joshua B. Oaks, MD (Progressive Clinical Research, Bountiful, UT)
22. Arin Piramzadian, DO (OnSite Clinical Solutions, Charlotte, NC)
23. Joe E. Pouzar Jr., MD (Centex Studies-Houston, Houston, TX)
24. Michael J. Rankin, MD (Worthington Urgent Care, Worthington, OH)
25. Ramon Reyes, MD (BFHC Research, San Antonio, TX)
26. Patricia D. Salvato, MD (Diversified Medical Practices, Houston, TX)
27. Jodi Sanson, MD (HealthStar Research, Hot Springs, AR)
28. Pantea Shoja, MD (Pearl City Urgent Care, Pearl City, HI)
29. Javier Sosa, MD (Hospital San Cristobal, Ponce, PR)
30. Alan Tannenbaum, MD (Vanguard Clinical Research, LLC, Fort Myers, FL)
31. Rafaelito Victoria, MD (Atella Clinical Research, La Palma, CA)
32. Kishor Vora, MD (Research Integrity, LLC, Owensboro, KY)
33. George S. Walker, MD (Best Clinical Trials, New Orleans, LA)
34. David Wever, MD (Cahaba Research-Pelham, Pelham, AL)
35. Michael Yuryev, DO (Integrative Clinical Trials, LLC, Brooklyn, NY)
36. Jeffrey Zacher, MD (West Valley Research Clinic, Phoenix, AZ)

### Supplementary Material

#### Definition of Subjects at Risk of Severe Illness (per CDC)

Subjects who are ≥65 years of age; subjects with COPD, Type 2 diabetes mellitus, obesity (BMI ≥30), chronic kidney disease, sickle cell disease, serious heart conditions (such as heart failure, coronary artery disease, or cardiomyopathies), asthma (moderate or severe), cerebrovascular disease, cystic fibrosis, hypertension or high blood pressure, immunocompromised state (due to immune deficiencies, HIV, use of corticosteroids, or use of other immune-weakening medications), neurologic conditions (e.g., dementia), liver disease, pulmonary fibrosis, past or present history of smoking, thalassemia, or type 1 diabetes mellitus.

