## Supplemental Material for "Early treatment with nitazoxanide prevents worsening of mild and moderate COVID-19 and subsequent hospitalization" (from another version of this preprint: version 1, 2021-04-20)

**Supplementary Material**

**Inclusion Criteria**

1. Male or female outpatients at least 12 years of age
2. Presence of clinical signs and/or symptoms consistent with worsening or stable mild or moderate COVID-19 (one of the following is required):
   a. Presence of at least two respiratory symptom domains (head, throat, nose, chest, cough) with a score of ≥2 as determined by Screening FLU-PRO© OR
   b. Presence of at least one respiratory symptom domain (head, throat, nose, chest, cough) with a score of ≥2 as determined by Screening FLU-PRO© with pulse rate ≥90 OR
   c. Presence of at least one respiratory symptom domain (head, throat, nose, chest, cough) with a score of ≥2 as determined by Screening FLU-PRO© with respiratory rate ≥16

   AND patient reported assessment that symptoms are present, the symptoms are not consistent with the subject's usual health, the symptoms interfere with daily activities, and the symptoms have worsened or remained the same relative to the previous day, as confirmed by responses to questions in the Screening FLU-PRO©.
3. Onset of symptoms no more than 72 hours before enrollment in the trial. Onset of symptoms is defined as the earlier of the first time at which the subject experienced subjective fever or any respiratory symptom (head, throat, nose, chest, or cough symptoms).
4. Willing and able to provide written informed consent (including assent by legal guardian if under 18 years of age) and comply with the requirements of the protocol, including completion of the subject diary and all protocol procedures.

**Exclusion Criteria**

1. Persons with any clinical sign or symptoms suggestive of severe systemic illness with COVID-19, including the following:
   a. shortness of breath at rest,
   b. resting pulse ≥125 beats per minute,
   c. resting respiratory rate ≥30 breaths per minute, or
   d. SpO2 ≤ 93% on room air at sea level.
2. Subjects who experienced a previous episode of acute upper respiratory tract infection, otitis, bronchitis or sinusitis or received antibiotics for these conditions within two weeks prior to and including study day 1.
3. Severely immunodeficient persons including:
   a. Subjects with immunologic disorders or receiving immunosuppressive therapy (e.g., for organ or bone marrow transplants, immunomodulatory therapies for certain autoimmune diseases)
   b. Subjects with untreated HIV infection or treated HIV infection with a CD4 count below 350 cells/mm3 in the last six months

   c. Subjects actively undergoing systemic chemotherapy or radiotherapy treatment for malignancy
   d. Subjects using steroids as maintenance therapy for chronic conditions
4. Subjects with active respiratory allergies or subjects expected to require anti-allergy medications during the study period for respiratory allergies.
5. Females of childbearing potential who are either pregnant or sexually active without the use of birth control. Female subjects of child-bearing potential that are sexually active must have a negative baseline pregnancy test and must agree to continue an acceptable method of birth control for the duration of the study and for 1 month post-treatment. A double barrier method, oral birth control pills administered for at least 2 monthly cycles prior to study drug administration, an IUD, or medroxyprogesterone acetate administered intramuscularly for a minimum of one month prior to study drug administration are acceptable methods of birth control for inclusion into the study. Female subjects are considered of childbearing potential unless they are postmenopausal (absence of menstrual bleeding for 1 year - or 6 months if laboratory confirmation of hormonal status), or have had a hysterectomy, bilateral tubular ligation or bilateral oophorectomy.
6. Subjects with a history of COVID-19 or known to have developed anti-SARS-CoV-2 antibodies.
7. Subjects residing in the same household with another subject participating in the study.
8. Treatment with any investigational drug or vaccine therapy within 30 days prior to screening and willing to avoid them during the course of the study.
9. Receipt of any dose of nitazoxanide within seven days prior to screening.
10. Known sensitivity to nitazoxanide or any of the excipients comprising the study medication.
11. Subjects unable to swallow oral tablets or capsules.
12. Subjects with known severe heart, lung, neurological or other systemic disease that the Investigator believes could preclude safe participation.
13. Subjects likely or expected to require hospitalization unrelated to COVID-19 during the study period.
14. Subjects taking medications considered to be major CYP2C8 substrates.
15. Subjects who, in the judgment of the Investigator, will be unlikely to comply with the requirements of this protocol including completion of the subject diary.

**Definition of Subjects at Risk of Severe Illness (per CDC):**
Subjects who are ≥65 years of age; subjects with COPD, Type 2 diabetes mellitus, obesity (BMI ≥30), chronic kidney disease, sickle cell disease, serious heart conditions (such as heart failure, coronary artery disease, or cardiomyopathies), asthma (moderate or severe), cerebrovascular disease, cystic fibrosis, hypertension or high blood pressure, immunocompromised state (due to immune deficiencies, HIV, use of corticosteroids, or use of other immune-weakening medications), neurologic conditions (e.g., dementia), liver disease, pulmonary fibrosis, past or present history of smoking, thalassemia, or type 1 diabetes mellitus.

**Supplementary Figure 1: Time-to-Event Analyses for Subjects in the Mild Illness Stratum at Baseline**

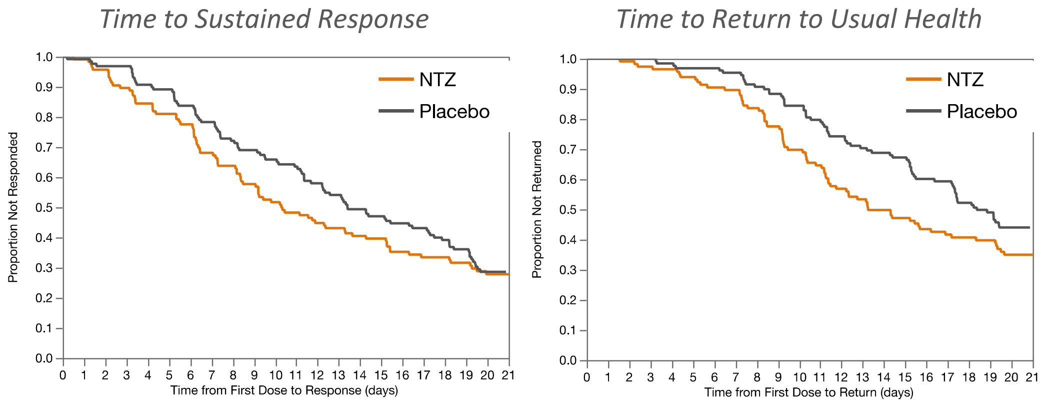
